# Life expectancy, quality of life, and hope among Japanese patients receiving home medical care

**DOI:** 10.1101/2022.09.13.22279424

**Authors:** Masakazu Yasunaka, Yukio Tsugihashi, Shinu Hayashi, Hidekazu Iida, Misaki Hirose, Yutaka Shirahige, Noriaki Kurita, the ZEVIOUS group

## Abstract

**Background:** Spiritual care should be included in home medical care for patients with limited life expectancy. However, the effect of shortened life expectancy on patients’ quality of life (QOL) and hopes is poorly understood.

**Methods:** This multicenter cross-sectional study involved 29 home medical care centers in Japan. Exposures were life expectancy (≥ one year / ≥ six months to < one year / < six months) as assessed by home medical care physicians. The outcomes were QOL in home medical care measured via the Quality-of-Life Scale for Elderly Patients Receiving Professional Home Care (QOL-HC), the domain scores of health-related hope (“health,” “role and connectedness,” and “something to live for”), and decrease in life functioning measured using the World Health Organization Disability Assessment Schedule 2.0 (WHODAS 2.0). Linear mixed models were fitted for analyses, with the facilities treated as clusters.

**Results:** Shorter life expectancy was associated with higher WHODAS 2.0 scores and significantly lower “something to live for” scores (< six months vs. ≥ one year). In contrast, “role and connectedness” scores did not change remarkably with decreased life expectancy, whereas QOL-HC scores were significantly higher with shorter life expectancy.

**Conclusion:** Home medical care physicians who engage in spiritual care should facilitate thoughtful dialogue with their patients by recognizing declines in life functions and hope for fulfillment, which are associated with short life expectancy.

## INTRODUCTION

Patients in Japan who require home medical care have impaired physical functions that render them unable to visit physicians (1–4). The leading causes vary, and include difficult-to-cure conditions such as cancer, multiple comorbidities including dementia and cerebrovascular disease, and neuromuscular diseases with severe disabilities (2, 5, 6). Therefore, when providing care to these patients, their lives at home should be supported in accordance with their preferences and needs (4, 7, 8). Other priorities include the maintenance of the quality of life (QOL), covering social and role functioning (3, 9), and relieving psychological distress (7). For patients with progressive illnesses, hope is a psychological state considered an inner resource and essential coping strategy for maintaining QOL (10). Coping and fostering hope are also considered central to clinical practice in palliative care (11). However, it remains unclear how hope and QOL evolve for patients receiving home medical care who typically desire to stay home for as long as possible until the end of their lives.

Hope involves the discovery of meaning to life (12) and the pursuit of future goals; moreover, along with QOL, it is closely associated with the well-being of patients with limited life expectancy (13–15). For example, the following question from a patient with advanced cancer—”Doctor, is there a chance of cure?”—is connected to hope regarding a cure (16). Furthermore, medical care providers are concerned that explicitly informing the patient of the lack of curative treatment and inevitable death may cause despair (8, 17). Although hope is essential for patients with advanced cancer, and multiple studies have reported an association between the progression of cancer and loss of hope (18–21), the association between remaining life expectancy and hope has not been investigated in home medical settings.

Providing goal-oriented care aligned with patients’ preferences and needs is expected to improve QOL among older patients with terminal cancer and advanced heart failure (10,22). It has also been emphasized as the benchmark for long-term care provided for older patients (3,9,23). However, few studies have examined the association between limited life expectancy, QOL, and hope in diverse populations receiving home medical care (some of whom are expected to have long life expectancies, regardless of severe disabilities, while others have limited life expectancy). Clarifying these associations enables home medical care physicians, who are in a unique position to directly provide palliative care (24, 25), to discuss the goals and values of care with their patients to maintain hope and QOL.

Therefore, the present study aimed to analyze the associations between expected life expectancy, QOL, and hope using data from a multicenter cross-sectional study conducted among patients receiving home medical care in Japan.

## METHODS

This study was conducted as part of the Zaitaku Evaluative Initiatives and Outcome Study (ZEVIOUS), which is a multicenter cross-sectional survey conducted in Japan between January and July 2020. Twenty nine home medical care facilities in the Tokyo Metropolitan area, Nara, and Nagasaki Prefectures were included in the study. The patients who consented to participate in the study were receiving continuous home medical care from home care physicians at the participating facilities, and were deemed by their physician as able to complete the questionnaire survey. The patients completed the questionnaire at their residence. Patients unable to write due to visual or physical impairments were allowed to complete the form with help from a family member or formal caregiver. The forms were mailed directly to the central research office to ensure that the completed questionnaires were not seen by the physicians treating the patients. This study was approved by the Institutional Review Board of Fukushima Medical University.

### Exposure

The home medicine physician assigned to the patient answered the following question: “How long do you expect the clinical prognosis (life expectancy) of this patient to be?” The physician was allowed to choose from five options: “less than one month,” “more than one month to less than three months,” “more than three months to less than six months,” “more than six months to less than 12 months,” and “more than 12 months.” Considering the frequency of the obtained responses, we merged them into the following three categories during the analysis phase: “less than six months,” “more than six months to less than 12 months,” and “more than 12 months.” These responses also considered a question proposed in a previous study about initiating a discussion about end-of-life care needs and preferences in the United Kingdom; it enquired whether the physician would be surprised if a patient were to die within a specific amount of time, defined as six months or one year (26).

### Outcomes

The following three constructs were measured as the outcomes in this study: the Health-Related Hope Scale (HR-Hope), Quality-of-Life Scale for Elderly Patients Receiving Professional Home Care (QOL-HC), and World Health Organization Disability Assessment Schedule 2.0 (WHODAS 2.0).

1. The HR-Hope Scale: This 18-item uni-dimensional scale assesses hope related to health among persons with chronic conditions (27). Through structural validation, the following three subdomains are scored: “something to live for” (five items), “health and illness” (six items), and “role and connectedness” (seven items). Responses to each item are rated on a four-point Likert scale ranging from 1 = *I don’t feel that way at all* to 4 = *I strongly feel that way*. After obtaining the average score for the total domain and each subdomain, the scores were scaled from 0 to 100. Patients without family were exempted from answering two items in the “role and connectedness” subdomain. The scale has been demonstrated to have sufficient internal consistency reliability (total domain, *“something to live for,” “health and illness,”* and *“role and connectedness”*: 0.93, 0.86, 0.86, and 0.74, respectively), criterion validity, and construct validity.
2. The QOL-HC: This four-item questionnaire assesses the QOL of older patients receiving home medical care (3). The face validity of the QOL-HC was ensured through the derivation of items by physicians and care managers and deliberate item selection by geriatricians (Supplementary Table 1). Each item is rated on a 3-point scale ranging from *“never agree”* (0 points) to *“always agree”* (2 points), with a total score ranging from 0 to 8 points. The internal consistency reliability (coefficient alpha = 0.7) and construct validity of the scale have been confirmed.

**Table 1.** Characteristics of patients separated by expected prognosis^*^ (n= 199)
3. The WHODAS 2.0: This 12-item scale measures functioning and disability, regardless of the disorders that cause dysfunction or disability, while complying with the International Classification of Functioning, Disability, and Health principles (28). It includes the following six domains: knowledge and communication (cognition), mobility, self-care, getting along with people (socializing), daily activities, and engagement in society. In addition, this scale includes items on challenges experienced over the past 30 days. The Japanese version of the 12-item WHODAS 2.0 has demonstrated excellent internal consistency reliability (coefficient alpha = 0.93) (29). Each item is scored on a 5-point scale ranging from *“none”* (1 point) to *“extreme/cannot”* (5 points), with the total score for each item converted to a scale of 0 to 100.

|  | Expected prognosis |  |  | Total<br>n = 199 |
| --- | --- | --- | --- | --- |
|  | ≥12mo | ≥6-<12mo | <6mo |  |
|  | n = 163 | n = 24 | n = 12 |  |
| **<u>Demographics</u>** |  |  |  |  |
| Age, yr | 79.8 (14.5) | 81.5 (12.5) | 80.5 (9.6) | 80 (14) |
| Women, n (%) | 92 (56.4 %) | 15 (62.5 %) | 10 (83.3 %) | 117 (58.8 %) |
| **Education, n (%)** |  |  |  |  |
| *Elementary school or junior high school* | 57 (35.6 %) | 7 (30.4 %) | 4 (33.3 %) | 68 (34.9 %) |
| *High school* | 51 (31.9 %) | 8 (34.8 %) | 4 (33.3 %) | 63 (32.3 %) |
| *College, university, or graduate school* | 52 (32.5 %) | 8 (34.8 %) | 4 (33.3 %) | 64 (32.8 %) |
| *missing, n* | 3 | 1 |  | 4 |
| Presence of family, n (%) | 143 (87.7 %) | 20 (83.3 %) | 12 (100 %) | 175 (87.9 %) |
| **Comorbidities, n (%)** |  |  |  |  |
| Cerebrovascular disease | 30 (18.4 %) | 3 (12.5 %) | 1 (8.3 %) | 34 (17.1 %) |
| Heart disease | 46 (28.2 %) | 11 (45.8 %) | 3 (25 %) | 60 (30.2 %) |
| Malignancy | 12 (7.4 %) | 8 (33.3 %) | 7 (58.3 %) | 27 (13.6 %) |
| Respiratory disease | 25 (15.3 %) | 6 (25 %) | 3 (25 %) | 34 (17.1 %) |
| Articular disease | 26 (16 %) | 1 (4.2 %) | 0 (0 %) | 27 (13.6 %) |
| Dementia | 34 (20.9 %) | 4 (16.7 %) | 0 (0 %) | 38 (19.1 %) |
| Neuromuscular disease | 22 (13.5 %) | 1 (4.2 %) | 0 (0 %) | 23 (11.6 %) |
| Fracture/Fall | 17 (10.4 %) | 3 (12.5 %) | 0 (0 %) | 20 (10.1 %) |
| Weakness | 27 (16.6 %) | 9 (37.5 %) | 1 (8.3 %) | 37 (18.6 %) |
| Spinal cord injury | 7 (4.3 %) | 0 (0 %) | 0 (0 %) | 7 (3.5 %) |

\*Means (SD) are presented for continuous data
Weakness: weakness associated with advancing age

### Other variables

Age, sex, educational attainment, presence of family members, and comorbidities were also recorded. The attending physician was asked to provide the comorbidities resulting in home medical care. Multiple choices were allowed, and other variables were collected using a patient questionnaire.

### Statistical analysis

Statistical analyses were performed using Stata/SE version 15. (StataCorp, College Station, TX, USA). Patient characteristics were described by means and standard deviations for continuous variables, and by frequencies and percentages for categorical variables for the overall and expected prognoses. The WHODAS 2.0, QOL-HC, and HR-Hope scores were similarly described for overall and expected prognoses. Mixed-effects linear regression models were performed with consideration of clustering effects by facility to evaluate the relationships between expected prognosis and the WHODAS 2.0, QOL-HC, and HR-Hope scores. For the QOL-HC analysis, we used robust variance estimation because the scale did not meet the standard assumptions of equal variances and normality. Age, sex, educational attainment, family, and comorbidities were entered into the models as covariates. In addition to the general linear models, other models were constructed by treating the expected prognosis as a continuous instead of a categorical variable to test the trends of monotone relationships between the expected prognosis and outcome variables (30).

Missing covariates were imputed using a multiple-imputation approach with chained equations. Statistical significance was defined as *p* < 0.05.

## RESULTS

Of the 202 patients receiving home medical care examined in this study, we excluded one patient without an expected prognosis and two patients who did not provide data for all three outcome variables. Consequently, data from 199 patients were used to investigate the associations between the expected prognosis and the WHODAS 2.0, QOL-HC, and HR-Hope scores (Figure 1). Table 1 presents patient characteristics. The mean age (standard deviation) was 80.0 (14) years; 117 (58.8%) of the participants were women. The comorbidities for which home medical care was required varied. Notably, cerebrovascular disease (*n* = 30, 18.4%), articular disease (*n* = 26, 16%), dementia (*n* = 34, 20.9%), neuromuscular disease (*n* = 22, 13.5%), and spinal cord injury (*n* = 7, 4.3%) were the most common among patients with expected prognoses of ≥ 12 months, and malignancy was the most common (*n* = 7, 58%) among patients with expected prognoses of < six months.

**Figure 1.**
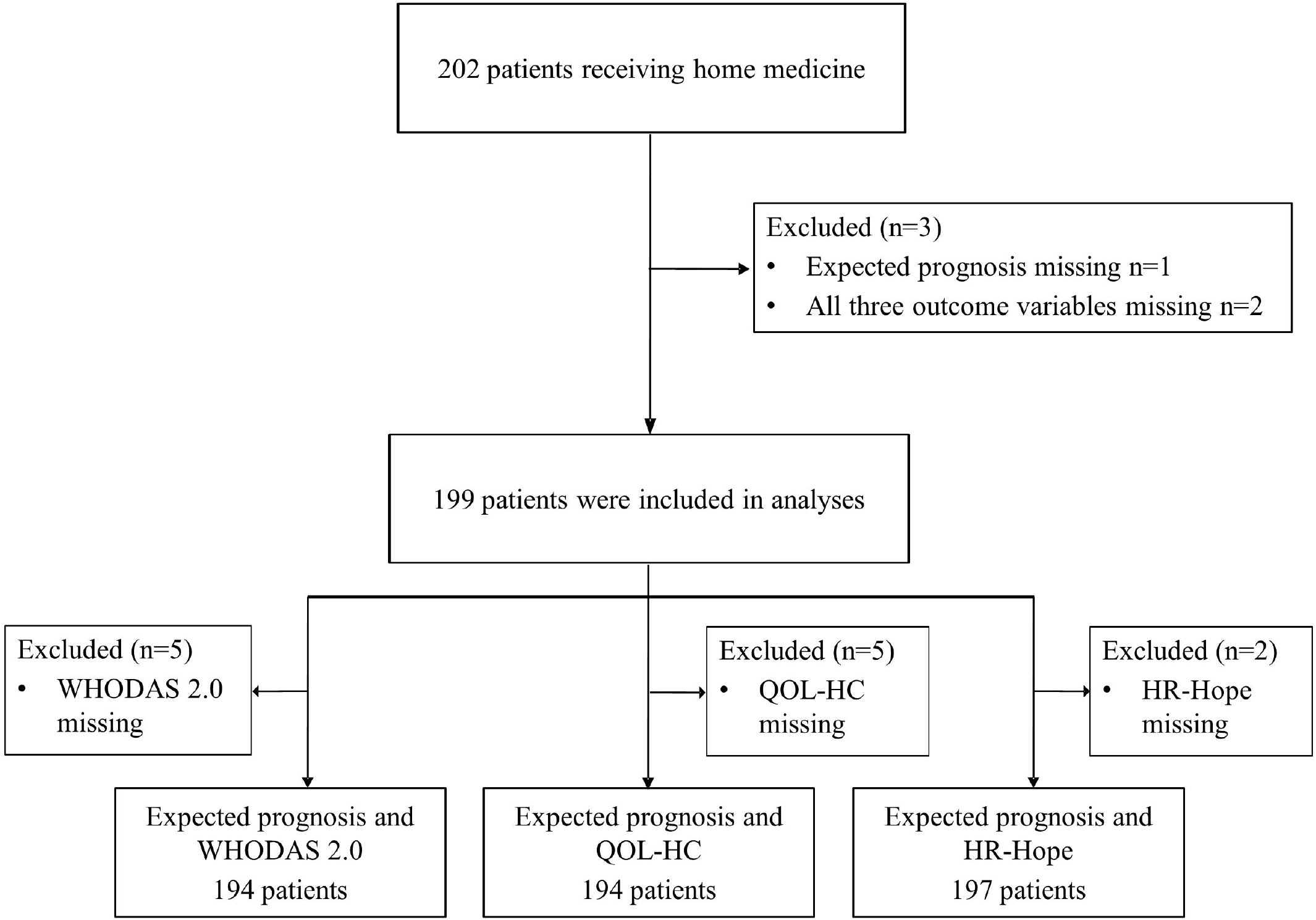
Flow diagram of the participants.

Table 2 shows the summary statistics for the WHODAS 2.0, QOL-HC, and HR-Hope scores for the overall patient population and expected prognosis subgroups. The means of the WHODAS 2.0 and QOL-HC scores among patients with an expected prognosis of < six months were higher than among those with an expected prognosis of ≥ 12 months. In contrast, the means of the HR-Hope overall and subdomain scores were lower among patients with an expected prognosis of ≥ 12 months.

**Table 2.** Description of outcomes separated by expected prognosis^*^ (n= 199)

|  | Expected prognosis |  |  | Total |
| --- | --- | --- | --- | --- |
| | $\geq 12mo$<br>n = 163 | $\geq 6- < 12mo$<br>n = 24 | $< 6mo$<br>n = 12 | |
| **WHODAS 2.0, pts** | 51.1 (25.4) | 54.5 (29.2) | 65.1 (31.2) | 52.3 (26.4) |
| *missing, n* | 4 | 1 |  | 5 |
| **QOL-HC, pts** | 6.4 (1.5) | 6.6 (1.2) | 6.9 (1.3) | 6.4 (1.4) |
| *missing, n* | 5 |  |  | 5 |
| **HR-Hope** |  |  |  |  |
| **Total, pts** | 57.6 (22.6) | 59.7 (24.9) | 48.3 (25.7) | 57.3 (23.1) |
| **Something to live for domain, pts** | 57.2 (26.2) | 58.9 (29.6) | 41.7 (27) | 56.5 (26.8) |
| *missing, n* | 1 |  |  | 1 |
| **Health domain, pts** | 53.7 (27) | 59.5 (23.6) | 43.1 (31.5) | 53.7 (27) |
| *missing, n* | 1 | 1 |  | 2 |
| **Connectedness domain, pts** | 60.7 (21) | 62.8 (24) | 57.5 (21.5) | 60.8 (21.4) |

\* Means (SD) are presented for continuous data
WHODAS 2.0: WHO Disability Assessment Schedule 2.0, HR-Hope: health-related hope, QOL-HC: QOL for patients receiving home-based medical care

Figure 2A and Supplementary Table 2 present the association between expected prognosis and WHODAS 2.0 scores. Compared to expected prognoses of ≥ 12 months, those of six to 12 months and < six months were associated with higher WHODAS 2.0 scores (13.9 [95%CI 2.5 - 25.3] and 19.6 points [95%CI 4.3 - 34.8], respectively). In addition, an increasing trend was observed in the WHODAS 2.0 scores as the expected prognosis decreased (*p* for trend = 0.002). Figure 2B and Supplementary Table 3 present the association between the expected prognosis and QOL-HC scores. Compared to expected prognoses of ≥ 12 months, those of < six months were associated with higher QOL-HC scores (0.7 points [95%CI 0.1 - 1.3]); expected prognoses of six to 12 months were not statistically associated with greater QOL-HC scores (0.3 points [95%CI -0.04 - 0.7]). However, an increasing trend was observed in QOL-HC scores as the expected prognosis decreased (*p* for trend = 0.006). Figure 2C and Supplementary Table 4 present the associations between the expected prognosis and HR-Hope subdomain scores. Compared to expected prognoses of ≥ 12 months, those of < six months were associated with lower “something to live for” scores (−17.7 points [95%CI -34.2 to -1.2]). However, evidence of a decreasing trend in the “something to live for” scores as the expected prognosis decreased was insufficient (*p* for trend = 0.074). Compared to expected prognoses of ≥ 12 months, evidence that those of < six months were associated with lower “health and illness” scores (−13.5 points [95%CI -30.5 to 3.5]) was insufficient. Additionally, evidence for a decreasing trend in the “health and illness” scores as the expected prognosis decreased (*p* for trend = 0.229) was insufficient. Compared to expected prognoses of ≥ 12 months, evidence that those of six to 12 months and < six months were associated with lower “role and connectedness” scores was insufficient (2.8 points [95%CI - 6.9 to 12.5] and -3.3 points [95%CI -16.4 to 9.8], respectively). Furthermore, evidence for a decreasing trend in the “role and connectedness” scores as the expected prognosis decreased was insufficient (*p* for trend = 0.901).

**Figure 2.**
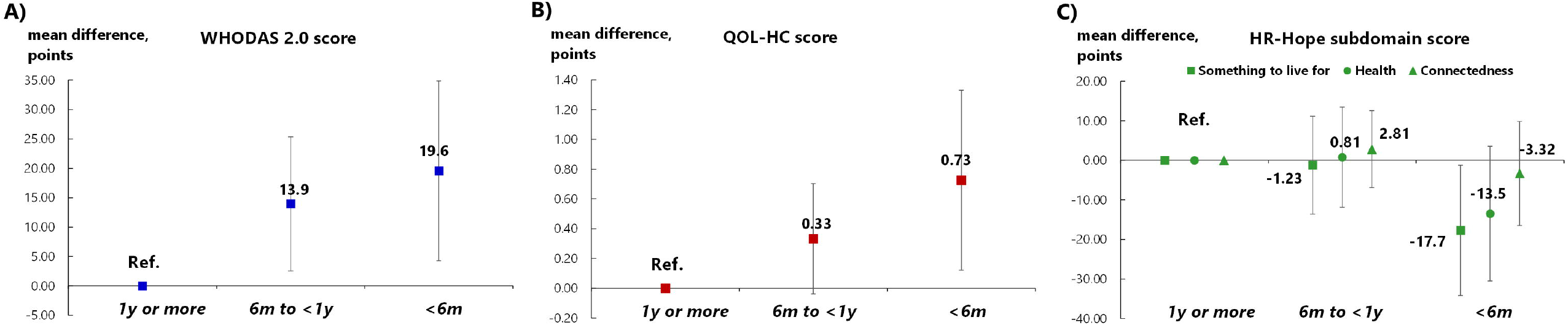
Associations between life expectancy and living function, quality of life, and health-related hope. A. Association between life expectancy and living function measured via WHODAS 2.0: Blue squares represent point estimates. B. Association between life expectancy and quality of life for home medical care measured by QOL-HC: Red squares represent point estimates. C. Associations between life expectancy and hope measured via the Health-Related Hope Scale:Green squares, circles, and triangles represent point estimates for something to live for, health, and role and connectedness, respectively. Estimates were derived from linear mixed models including age, sex, educational attainment, presence of family members, and comorbidities (cerebrovascular disease, heart disease, malignancy, respiratory disease, articular disease, dementia, neuromuscular disease, fractures or falls, weakness, spinal cord injury) as covariates.

## DISCUSSION

Among the patients receiving home medical care, short life expectancy was associated with lower hope for “something to live for,” although hope for “role and connectedness” was maintained. Additionally, short life expectancy was associated with higher QOL in relation to current home medical care, even though it was associated with decreased functioning.

The findings of the present study on the relationship between life expectancy and hope and QOL in home medical care are consistent with previous findings in the palliative care field, and provide exceptional results in home medical care. First, the lower “something to live for” domain scores among patients with reduced life expectancy were consistent with previous studies reporting decreased hope for the future following perceived physical deterioration (10, 17, 31). Although not statistically replicable, lower hope scores for health were associated with shorter life expectancy in our study sample. The present study confirms the presence of psycho-existential suffering, proposed in palliative care and in home medical care settings, combining our findings of lower perceived living function in patients with shorter life expectancy with the findings on hope. In other words, this study suggests that home medical care patients with reduced life expectancy may suffer from loss of autonomy (which includes physical independence, a sense of control over the future, and continuity of fulfillment in life) and temporality (which includes anxiety about death, recovery from illness, and hope for the achievement of goals) (32). Second, the finding about hope for “role and connectedness,” which did not decrease even within six months of life expectancy, may indicate a benefit of receiving home medical care. This idea is supported by the maintenance of “connectedness with others” by a high percentage (approximately 90%) of patients with family members in this study; family relationships are maintained through three-way communication between patients, their families, and home care physicians, and frequent visits by a multidisciplinary home medical care team (6). In other words, the loss of relationships—including the desire to be with the family and not be a burden to family members or health-care providers (32)—is minimized among patients receiving home medical care even if they have short life expectancies. Third, the unexpected relationship between short life expectancy and high QOL measured by the QOL-HC may be explained by the increased frequency of patients’ reflections on their life, which is part of this outcome measure. A report on improved QOL due to increased life-reviewing near the end of life partially explains our findings (33). Alternative explanations include improved patient satisfaction through interactions with home medical care providers and improved QOL caused by reduced pain and other symptoms through appropriate palliation at the end of life (8, 34).

This study has several implications for home medical care practitioners and researchers. First, home medical care providers should initiate dialogue with patients with limited life expectancy, while considering that, even if their QOL improved through home medical care, they may not necessarily maintain hope or may have lost some hope. For example, a question (e.g., “Doctor, do you think I will be cured?”) from patients with limited life expectancy can be interpreted as their reflection of wavering hope rather than genuine hope for recovery from their illness. Therefore, home medical care providers should confirm patients’ perceptions regarding their disease progression and responsiveness to treatment, and provide realistic explanations to fill the gap between their perceptions and the care providers’ evaluations (16). During this dialogue, the home care physician should share realistic goals of care (e.g., relieving symptoms and enjoying activities such as socializing with loved ones) with the patient, and explain how these goals are achievable through medical intervention (10). Such dialogue that encourages end-of-life hope without negatively impacting the patient may lead their family and them to perceive the end-of-life experience as good (35). Second, this study highlights the need for a psychosocial intervention for home medical care patients with short life expectancies who may suffer from a loss of autonomy and temporality due to the pursuit of unrealistic goals. Interventions that enable patients to shift from future-oriented thinking— such as continuity of fulfillment in life and achievement of long-term goals—to a life-review method that encourages reflection on past experiences that one is proud of contribute to maintaining hope in patients with advanced cancer and chronic illnesses (10, 33, 36).

The present study has several strengths. First, this study is the first to measure health-related hope by domain and correlate it with life expectancy among patients receiving home medical care. Second, our findings are generalizable because the study was conducted in a multicenter setting—urban and rural home medical care facilities—and the analyses accounted for differences in the clustering of outcomes across these facilities. Third, as indicated in a previous study (37), the study was designed so that the questionnaires were answered in the absence of the attending home medical care physician, and the forms were mailed directly to the central facility.

Nonetheless, this study has some limitations. First, this study was conducted among patients who could answer the questionnaire independently. Therefore, patients with severe dementia, reduced consciousness, or those who may die within a few days were not included. Second, perceptions regarding the HR-Hope scale may vary across countries. While cultural and religious backgrounds and perceptions of spirituality may differ between Asian and Western countries (18), racial differences in tolerance of religious hope and psychological distress have also been reported (38).

In summary, short life expectancy was associated with higher QOL in relation to home medical care and with lower functioning and hope for “something to live for” among Japanese patients receiving home medical care. This study highlights the urgent need for home physicians to deliver a good end-of-life experience by focusing on hope.

## Data Availability

All data produced in the present work are contained in the manuscript

## Ethical approval

This study was approved by the Institutional Review Board of Fukushima Medical University (ippan-30254).

## Acknowledgments

The authors express their sincere gratitude to the research assistants, Ms. Miyuki Sato and Ms. Lisa Shimokawa (Fukushima Medical University Hospital, Fukushima City, Fukushima), for their assistance in collecting the questionnaire-based information used in this study.

