## Supplementary Table1-4 and Text for "Life expectancy, quality of life, and hope among Japanese patients receiving home medical care"

**Supplementary Table 1.　QOL for patients receiving home-based medical care (QOL-HC).**

| Stem |  |
| --- | --- |
| Question 1 | Do you have peace of mind? |
| Question 2 | Do you feel satisﬁed with your life when you reﬂect on it? |
| Question 3 | Do you have someone that you spend time talking with? |
| Question 4 | Are you satisﬁed with the home care service system? |
| Response options for Question | Never agree / Neither agree nor disagree/ always agree |

The original English version is also provided for each item and response.

### **Supplementary Table 2. Associations between prognostic expectation and WHODAS 2.0**^*^ **(n = 194)**

| WHODAS 2.0, points | mean difference (95%CI) | P-value |
| --- | --- | --- |
| Expected prognosis |  |  |
| ≥ 12 month | Reference |  |
| ≥ **6 - < 12 month** | **13.9 (2.5 - 25.3)** | **0.017** |
| **< 6month** | **19.6 (4.3 - 34.8)** | **0.012** |
| **Age, per 10y** | **3.3 (0.5 - 6.1)** | **0.021** |
| Women vs. Men | 2.2 (-5.03 - 9.4) | 0.553 |
| Educational attainment |  |  |
| Junior high school or lower | Reference |  |
| High school | -5.2 (-13.7 - 3.3) | 0.232 |
| College/University/Graduate school/Other | 0.1 (-8.5 - 8.8) | 0.973 |
| **Presence of family** | **11.3 (1.2 - 21.3)** | **0.028** |
| Comorbidities |  |  |
| **Cerebrovascular disease** | **16.02 (6.6 - 25.4)** | **0.001** |
| Heart disease | -6.1 (-13.9 - 1.6) | 0.122 |
| Malignancy | -11.04 (-22.8 - 0.7) | 0.066 |
| Respiratory disease | 7.3 (-1.6 - 16.3) | 0.109 |
| Articular disease | 9.3 (-0.8 - 19.4) | 0.071 |
| Dementia | 4.5 (-4.9 - 13.9) | 0.347 |
| **Neuromuscular disease** | **16.4 (4.8 - 28)** | **0.006** |
| Fracture/Fall | 5.7 (-5.5 - 16.9) | 0.321 |
| Weakness | 3.02 (-7.1 - 13.2) | 0.560 |
| **Spinal cord injury** | **22.7 (3.8 - 41.6)** | **0.018** |

Analysis of 194 patients among 29 facilities.

^*^Mixed-effects linear regression models adjusted for covariates listed above.

### **Supplementary Table 3. Associations between prognostic expectation and QOL-HC**^*^ **(n = 194)**

| QOL-HC, points | mean difference (95%CI) | P-value |
| --- | --- | --- |
| Expected prognosis |  |  |
| ≥ 12 month | Reference |  |
| ≥ 6 - < 12 month | 0.3 (-0.04 - 0.7) | 0.078 |
| **< 6 month** | **0.7 (0.1 - 1.3)** | **0.019** |
| Age, per 10y | 0.1 (-0.004 - 0.3) | 0.057 |
| Women vs. Men | 0.1 (-0.3 - 0.6) | 0.551 |
| Educational attainment |  |  |
| Junior high school or lower | Reference |  |
| High school | -0.18 (-0.7 - 0.3) | 0.486 |
| College/University/Graduate school/Other | -0.3 (-0.8 - 0.1) | 0.146 |
| Presence of family | 0.2 (-0.3 - 0.7) | 0.418 |
| Comorbidities |  |  |
| Cerebrovascular disease | 0.02 (-0.6 - 0.7) | 0.960 |
| Heart disease | 0.1 (-0.4 - 0.5) | 0.798 |
| Malignancy | -0.47 (-1.01 - 0.1) | 0.088 |
| Respiratory disease | 0.3 (-0.2 - 0.8) | 0.230 |
| Articular disease | -0.2 (-0.9 - 0.5) | 0.553 |
| **Dementia** | **0.6 (0.1 - 1.01)** | **0.015** |
| Neuromuscular disease | -0.2 (-0.98 - 0.5) | 0.528 |
| Fracture/Fall | -0.1 (-0.6 - 0.4) | 0.753 |
| Weakness | -0.14 (-0.6 - 0.3) | 0.558 |
| Spinal cord injury | 0.1 (-0.8 - 1.04) | 0.777 |

Analysis of 194 patients among 29 facilities.

^*^Mixed-effects linear regression models adjusted for covariates listed above with robust standard errors.

**Supplementary Table 4. Associations between expected prognosis, covariates, and HR-Hope domains**^*^ **(n = 197)**

|  | **Something to live for** |  | **Health and Illness** |  | **Role and connectedness** |
| --- | --- | --- | --- | --- | --- |
|  | mean difference, point estimate (95%CI) |  | mean difference, point estimate (95%CI) |  | mean difference, point estimate (95%CI) |
| Expected prognosis |  |  |  |  |  |
| ≥ 12 month | Reference |  | Reference |  | Reference |
| ≥ 6 - < 12 month | -1.2 (-13.6 to 11.1), p = 0.846 |  | 0.8 (-11.8 to 13.5), p = 0.9 |  | 2.8 (-6.9 to 12.5), p = 0.571 |
| < 6 month | **-17.7 (-34.2 to -1.2), p = 0.035** |  | -13.5 (-30.5 to 3.5), p = 0.12 |  | -3.3 (-16.4 to 9.8), p = 0.62 |
| Age, per 10y | -1.2 (-4.2 to 1.8), p = 0.433 |  | -2.4 (-5.5 to 0.7), p = 0.135 |  | -1.4 (-3.8 to 0.999), p = 0.25 |
| Women vs. Men | -0.96 (-8.8 to 6.9), p = 0.809 |  | 0.5 (-7.7 to 8.6), p = 0.91 |  | 1.6 (-4.7 to 7.9), p = 0.622 |
| Educational attainment |  |  |  |  |  |
| Junior high school or lower | -3.4 (-12.5 to 5.8), p = 0.468 |  | -4.02 (-13.5 to 5.5), p = 0.406 |  | -1.3 (-8.6 to 5.9), p = 0.716 |
| High school | **-11.3 (-20.6 to -2), p = 0.018** |  | -9.6 (-19.3 to 0.1), p = 0.052 |  | -6.5 (-13.9 to 0.998), p = 0.09 |
| College/University/Graduate school/Other | Reference |  | Reference |  | Reference |
| Presence of family | -5.04 (-15.8 to 5.7), p = 0.359 |  | -5.4 (-16.6 to 5.8), p = 0.345 |  | 6.4 (-2.3 to 15), p = 0.148 |
| Comorbidities |  |  |  |  |  |
| Cerebrovascular disease | -4.6 (-14.7 to 5.6), p = 0.377 |  | -5.01 (-15.5 to 5.5), p = 0.35 |  | **-9.2 (-17.3 to -1.04), p = 0.027** |
| Heart disease | 5.8 (-2.5 to 14), p = 0.172 |  | 3.6 (-5 to 12.1), p = 0.415 |  | 3.6 (-3.03 to 10.2), p = 0.288 |
| Malignancy | 7.9 (-4.4 to 20.2), p = 0.21 |  | 4.1 (-8.6 to 16.7), p = 0.529 |  | 0.4 (-9.3 to 10.2), p = 0.93 |
| Respiratory disease | -8.3 (-18 to 1.3), p = 0.09 |  | -4.8 (-14.8 to 5.2), p = 0.348 |  | -4.2 (-11.9 to 3.5), p = 0.286 |
| Articular disease | -4.8 (-15.7 to 6), p = 0.381 |  | -8.5 (-19.7 to 2.6), p = 0.134 |  | -4.4 (-13 to 4.2), p = 0.319 |
| Dementia | 4.9 (-5.1 to 15), p = 0.336 |  | 1.1 (-9.2 to 11.4), p = 0.831 |  | 7.2 (-0.8 to 15.1), p = 0.077 |
| Neuromuscular disease | -3.7 (-16.3 to 8.9), p = 0.562 |  | -9.8 (-22.8 to 3.3), p = 0.142 |  | -1.8 (-11.9 to 8.3), p = 0.726 |
| Fracture/Fall | 5.8 (-6.4 to 18), p = 0.354 |  | 7.95 (-4.6 to 20.5), p = 0.215 |  | 3.6 (-6.1 to 13.3), p = 0.465 |
| Weakness | -1.6 (-12.6 to 9.3), p = 0.77 |  | -2.6 (-13.9 to 8.7), p = 0.653 |  | 2.4 (-6.3 to 11.1), p = 0.587 |
| Spinal cord injury | -12.2 (-32.8 to 8.3), p = 0.243 |  | -9.9 (-31.1 to 11.3), p = 0.36 |  | -2.1 (-18.5 to 14.3), p = 0.801 |

Analysis of 197 patients among 29 facilities.

^*^Mixed-effects linear regression models adjusted for covariates listed above.

**Supplementary Text. Full details of the other members of the ZEVIOUS Group**

Shinsuke Muto^1^; Tatsunobu Natsubori^1,2^; Michiko Hinata^1,3^; Wataru Nakagawa^1^; Akihiko Yonenaga^1^; Lina Inagaki^1^; Shioto Itakura^1^; Nobuhiro Ikeda^1^; Tomoka Nakamura^1^; Naoya Miyashita^1^; Takuya Furugen^1^; Takafumi Abo^4,5^; Sadayuki Okudaira^4,6^; Kazuhiko Takuma^4,7^; Chihiro Tsuchiya^,8^; Masahiro Deguchi^4,9^; Takashi Fujii^4,10^; Yoshitaka Harada^4,11^; Seiji Matsuo^4,12^; Motomichi Nakagawa^4,13^; Ken Tanigawa^4,14^; Yoshio Ochi^4,15^; Sadanobu Ogasawara^4,16^; Kazuhiko Hoshino^4,17^; Momoko Aruga^18^; Yoshinori Nakamura^18^; Nobuhiro Sawa^19^; Yosuke Akashi^19^; Nobuyuki Miyagi^20^; Toyohiro Terasaki^21^; Kunihiro Kinoshita^22^; Masaji Kikukawa^23^; Hisakazu Kato^24^; Masayuki Amano^25^; Kentaro Asakura^26^; and Naoto Fukui^27^.

^1^You Home Clinic, Bunkyo City, Japan

^2^You Home Clinic Azumabashi, Sumida City, Japan

^3^You Home Clinic Azabudai, Minato City, Japan

^4^Dr. Net Nagasaki, Nagasaki City, Japan

^5^Abo Gastrointestinal Surgical Clinic, Nagasaki City, Japan

^6^Okudaira Geka, Nagasaki City, Japan

^7^Takuma Clinic, Nagasaki City, Japan

^8^Chihiro Naika Clinic, Nagasaki City, Japan

^9^Deguchi Surgery Clinic, Nagasaki City, Japan

^10^Fujii Surgical Clinic, Nagasaki City, Japan

^11^Harada Internal Medicine Clinic, Nagasaki City, Japan

^12^Nagasaki Takara Home Medical Care Clinic, Nagasaki City, Japan

^13^Nakagawa Surgical Clinic, Nagasaki City, Japan

^14^Tanigawa Clinic, Nagasaki City, Japan

^15^Ochi Clinic, Nagasaki City, Japan

^16^Nagasaki Memorial Hospital, Nagasaki City, Japan

^17^Hoshino Internal and Respiratory Medical Clinic, Nagasaki City, Japan

^18^Medical home care center, Tenri Hospital Shirakawa Branch, Tenri City, Japan

^19^Minami Nara General Medical Center, Oyodo, Japan

^20^Miyagi Clinic, Tenri City, Japan

^21^Terasaki Clinic, Nara City, Japan

^22^Kinoshita Clinic, Sakurai City, Japan

^23^Kikukawa Internal Medicine Clinic, Sakurai City, Japan

^24^Kato Clinic, Uda City, Japan

^25^Nosegawa Village National Health Insurance Clinic, Nosegawa, Japan

^26^Daifuku Clinic, Sakurai City, Japan

^27^Fukui Clinic, Uda City, Japan
